# Cardiovascular magnetic resonance imaging after normal echocardiography in myocardial infarction with nonobstructed coronary arteries

**DOI:** 10.1101/2022.10.25.22281518

**Authors:** Martin G. Sundqvist, Peder Sörensson, Christina Ekenbäck, Magnus Lundin, Stefan Agewall, Elin Bacsovics Brolin, Kerstin Cederlund, Olov Collste, Maria Daniel, Jens Jensen, Shams Y-Hassan, Loghman Henareh, Claes Hofman-Bang, Patrik Lyngå, Eva Maret, Nondita Sarkar, Jonas Spaak, Oscar Winnberg, Kenneth Caidahl, Martin Ugander, Per Tornvall

## Abstract

**Background:** In patients with myocardial infarction with nonobstructive coronary arteries (MINOCA), cardiovascular magnetic resonance imaging (CMR) can often establish a causal diagnosis. It is unknown if CMR is warranted in patients with normal echocardiographic findings, or if measurement of high-sensitive troponin T (hs-TnT) and NT-pro-BNP can be of value in selecting patients for further imaging.

**Objectives:** To assess the diagnostic accuracy of echocardiography and hs-TnT and NT-pro-BNP for identifying patients receiving any diagnosis using CMR.

**Methods:** We included patients (n = 123) from the SMINC2 study who underwent same day CMR and echocardiography, at a median of 3 days after hospital admission for MINOCA. Normal echocardiography was defined as left ventricular ejection fraction ≥ 55%, absolute global longitudinal strain ≥ 17%, E/e′ ≤ 14, and no regional wall motion abnormalities. Logistic regression models were fitted to assess the probability of CMR diagnosis at increasing levels of hs-TnT and NT-pro-BNP.

**Results:** Of patients with a normal echocardiographic examination, 23/33 (70%) received a diagnosis using CMR. Pathological echocardiography identified patients with a diagnosis using CMR with a sensitivity of 77%, specificity 38%, positive predictive value 82%, and negative predictive value 30%, respectively. There was no level of hs-TnT or NT-pro-BNP below which a CMR diagnosis could be reliably excluded.

**Conclusions:** The majority of patients with MINOCA and a normal echocardiogram will receive a diagnosis by CMR. A CMR diagnosis was common even among patients with low levels of biomarkers. CMR should be recommended regardless of echocardiographic findings and hs-TnT or NT-pro-BNP levels in patients with MINOCA.

## Introduction

Myocardial infarction with nonobstructed coronary arteries (MINOCA) is an increasingly recognized clinical entity, constituting approximately 5-9% of all cases of myocardial infarction (MI)^1,2^. Being a working diagnosis, MINOCA warrants further investigation to establish a final diagnosis. In most patients, the final diagnosis is takotsubo syndrome (TTS), myocarditis, or MI, but any disease process temporarily mimicking an acute coronary syndrome could be implicated^3^. The increasing interest in MINOCA has led to a recognition of the value of cardiovascular magnetic resonance imaging (CMR) in establishing a definitive diagnosis^4,5^, which is successful in at least 75% of the cases^6,7^. Optical coherence tomography (OCT) of the coronary arteries has been proposed to increase the number of patients who receive a diagnosis^8,9^, but CMR is unique in its ability to not only visualize the consequences of ischemia such as myocardial edema and scarring in the territory of a coronary artery, but also, to detect various other conditions through a combination of anatomical and functional imaging, and myocardial tissue characterization.

TTS is arguably the most common diagnosis among patients with MINOCA as a working diagnosis, and patients with TTS have been shown to have higher plasma levels of NT-pro-BNP than patients with MI, in particular in relation to plasma troponin levels^10^. There is also a strong correlation between troponin T levels and MI size determined by CMR^11^. Hypothetically, it seems likely that the chance of obtaining a diagnosis using CMR is associated with the level of increase of plasma biomarkers.

To the best of our knowledge no study has examined the diagnostic value of echocardiography in MINOCA in a head-to-head comparison. In this study, we aimed to investigate to what extent patients with normal echocardiographic findings still would receive a diagnosis by CMR, and whether using plasma levels of hs-TnT and NT-pro-BNP could aid in this assessment. Our hypothesis was that patients with a normal echocardiography would have a normal CMR investigation.

## Methods

### Study group and CMR

The SMINC-2 study enrolled 148 patients with MINOCA, and a comprehensive CMR examination including T1- and extracellular volume fraction mapping, contrast-enhanced cine, and motion-corrected phase sensitive inversion recovery sequences for late gadolinium enhancement analysis was performed (median [interquartile range]) 3 [2–4] days after admission. Patients were classified into diagnosis groups based solely on CMR findings, as described previously^6^. In summary, 77% of all patients received a diagnosis using CMR, 35% had TTS, 22% MI and 17% had myocarditis. There were three cases of dilated cardiomyopathy and two cases of hypertrophic cardiomyopathy. As these diagnoses were distinct but rare, these five patients were excluded from further analysis, since this study focuses on the value of CMR in patients with MINOCA without a clear underlying cause. The SMINC2 study was performed in accordance with the Declaration of Helsinki and good clinical practice and was approved by the Stockholm Regional Board of Ethics (2014/131-31/1, 2014/1546-32). All patients provided written informed consent.

### Echocardiography and blood sampling

Echocardiography on the same day as CMR was available in 128 patients, as part of a planned sub-study. All echocardiography examinations were performed at Karolinska University Hospital by dedicated staff using Vivid E95 scanners (GE, Horten, Norway), in accordance with relevant clinical guidelines. A contrast agent (SonoVue, Bracco) was administered as part of the protocol, the contrast-enhanced images were used for evaluation of regional wall motion abnormalities (RWMA), but not ventricular volumes. All measurements and analyses were performed in EchoPAC v203 (GE, Horten, Norway) by one observer (MGS) without knowledge of the CMR diagnosis. Global longitudinal strain (GLS) was calculated as the mean of longitudinal strain from at least two apical views and reported as an absolute percentage. The presence of RWMA was assessed in a 16-segment model. Diastolic function was classified using the 2016 echocardiography guidelines^12^. A normal echocardiographic examination was defined as left ventricular ejection fraction (LVEF) ≥ 55% without RWMA, absolute global longitudinal strain ≥ 17%, and E/e′ ≤ 14. E/e′ was used as a proxy for diastolic function, as the classification method of the 2016 guidelines often leads to a substantial amount of indeterminate cases^13^. Plasma hs-TnT and NT-pro-BNP concentrations were analyzed by standard methods by the Karolinska university hospital laboratory in blood samples drawn on the same day as the CMR and echocardiographic examinations.

### Statistical analysis

Echocardiographic and laboratory findings, and demographic data for patients with and without a CMR diagnosis were compared with the Wilcoxon rank-sum test for continuous variables and the chi-square test for categorical variables. For the calculation of sensitivity, specificity, positive and negative predictive value (PPV and NPV, respectively), a positive CMR was defined as one establishing a diagnosis, and a pathological echocardiogram was defined as one not fulfilling the criteria for a normal echocardiogram. For the sake of investigating how the probability of obtaining a CMR diagnosis changes with increasing levels of hs-TnT and NT-pro-BNP, logistic regression models were fitted with the respective biomarkers and echocardiography findings divided into normal/pathological as dependent variables and presence of any CMR diagnosis as binary independent variable. The models were then used to predict the probability of any CMR diagnosis which was reported graphically, and as odds ratios with full model specifications reported in the Supplemental material. All statistical analyses were performed in R version 4.1.2 (R Core Team, 2021)^14^, regression modeling in rms^15^ and graphs produced using ggplot2^16^. A p-value of less than 0.05 was considered statistically significant.

## Results

Echocardiography was available in 123 patients. Echocardiography was assessed as normal in 33 cases and pathological in 90 cases.

### Diagnosis by CMR

According to the CMR results of the patients who had an available echocardiography, 47 patients (38%) had TTS, 27 (22%) had MI, 23 (19%) had myocarditis, and 26 (21%) had no identified causal diagnosis.

### Diagnostic accuracy of a normal echocardiographic examination

The results of the echocardiographic examinations are reported in Table 1, grouped by the presence or absence of a causal diagnosis by CMR. The only statistically significant difference was a higher number of hypokinetic segments (0 [0–3] vs 0 [0–1], p = 0.03) in the group with a final CMR diagnosis.

**Table 1.**
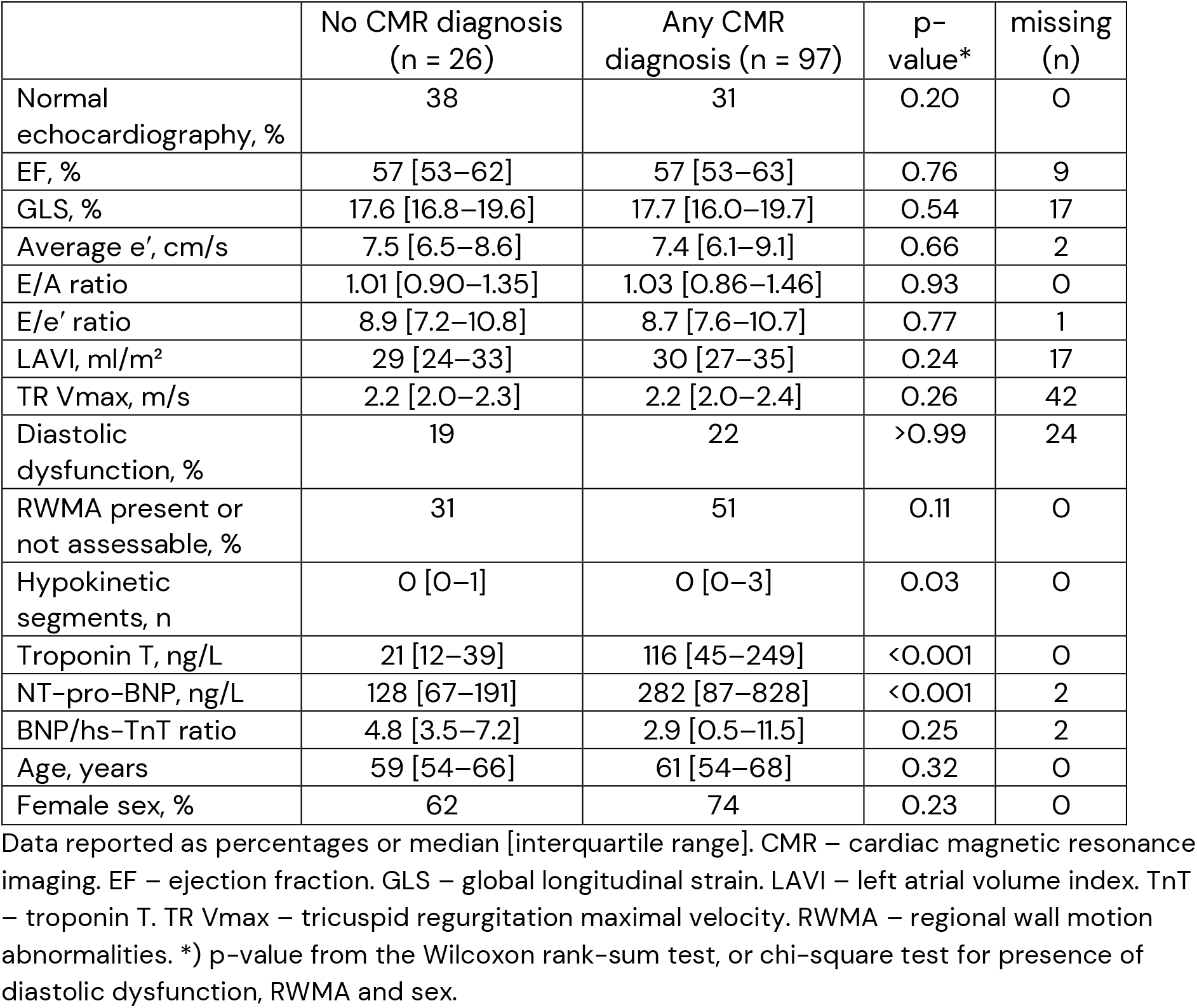
Echocardiographic and laboratory findings, and demographic data

Of all echocardiographic examinations, 33/123 (27%) were assessed as normal according to the criteria of this study. Of these, 23/33 (70%) received a diagnosis using CMR. The echocardiography criteria had sensitivity 76%, specificity 38%, PPV 82% and NPV 30%. The relationship between echocardiographic assessment and the different diagnosis groups is illustrated in the Central illustration. Of the patients with normal echocardiography, 3 had TTS, 10 MI and 10 had myocarditis. Of the patients with pathological echocardiography, 16 had non-diagnostic CMR findings.

### Probability of establishing a diagnosis using hs-TnT or NT-pro-BNP

Patients receiving a diagnosis by CMR had higher hs-TnT (116 [45–249] vs 21 [12–39] ng/L, p < 0.001) and NT-pro-BNP (282 [87–828] vs 128 [67–191] ng/L, p < 0.001) compared to those without diagnosis, but similar NT-pro-BNP/hs-TnT-ratio (p = 0.25). The distribution of biomarker concentrations on a logarithmic scale in the patients with and without a causal CMR diagnosis is illustrated in Figure 2. The distribution of diagnoses per tertile of respective biomarker level is illustrated in Figure 3. MI was a more common diagnosis with increasing levels of hs-TnT, TTS was respectively more common with increasing levels of NT-pro-BNP. The logistic regression model results are available in the supplemental material. The odds ratio for establishing a CMR diagnosis with a 10-unit increase in hs-TnT was 1.27 (95% confidence interval (CI) 1.11–1.46), and 1.45 (95% CI 1.08–1.94) with a 100-unit increase in NT-pro-BNP, respectively. The predicted probabilities, derived from these logistic regression models, of establishing any diagnosis are presented graphically in Figure 4. As can be seen, even at the lower limits of hs-TnT and NT-pro-BNP, the probability for achieving a diagnosis was predicted to be approximately 0.25 or 0.5, respectively. Thus, there was no level of hs-TnT or NT-pro-BNP below which a CMR diagnosis could be reliably excluded. The predicted probability tended to be stronger with a pathological than normal echocardiogram for hs-TnT but did not seem to differ for NT-pro-BNP.

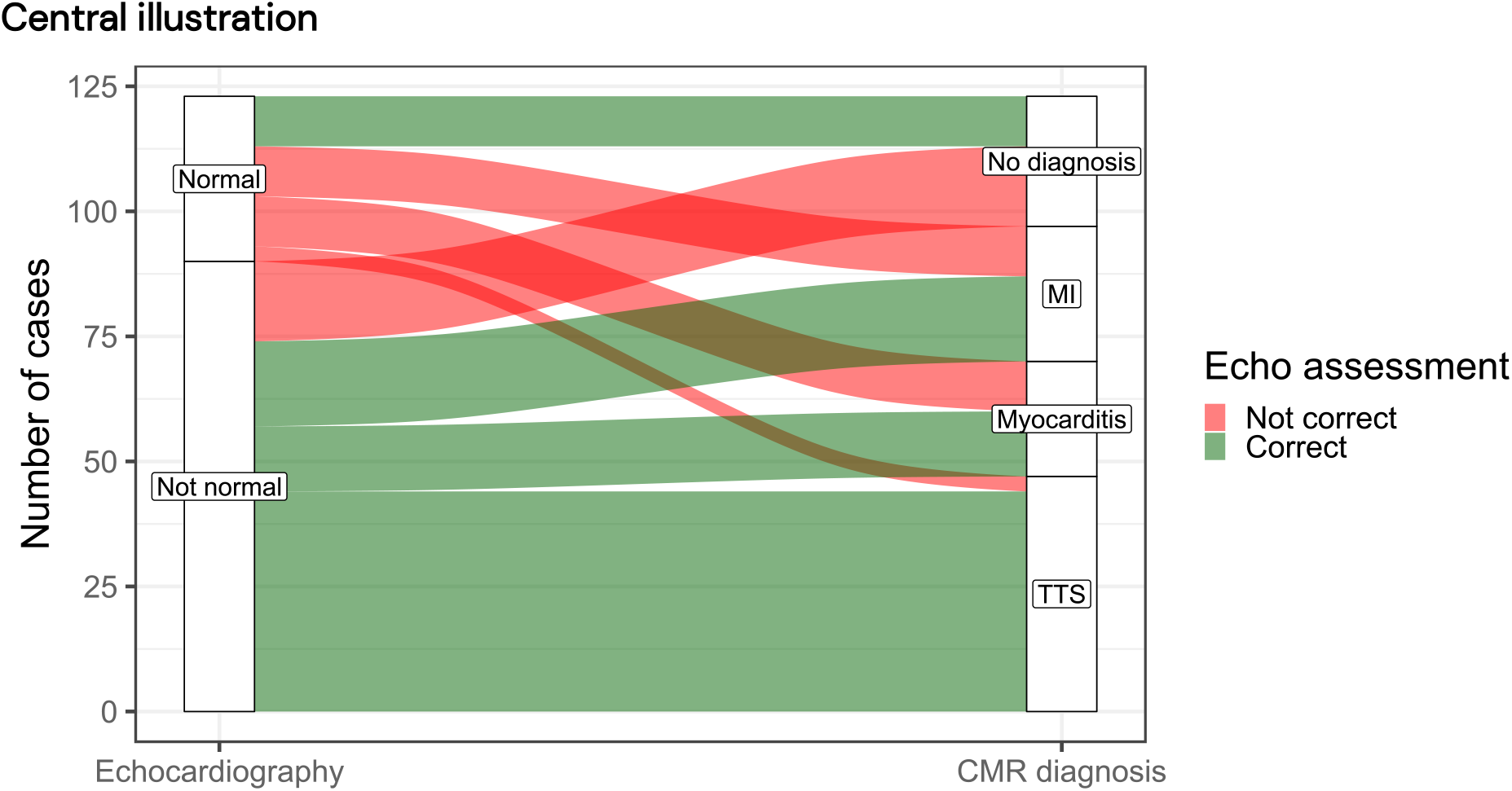
Alluvial plot illustrating the final cardiac magnetic resonance (CMR) diagnosis in patients with normal and pathological echocardiography. Red and green color indicates incorrect and correct classification by echocardiography, respectively. MI – myocardial infarction. TTS – takotsubo syndrome.

**Figure 2.**
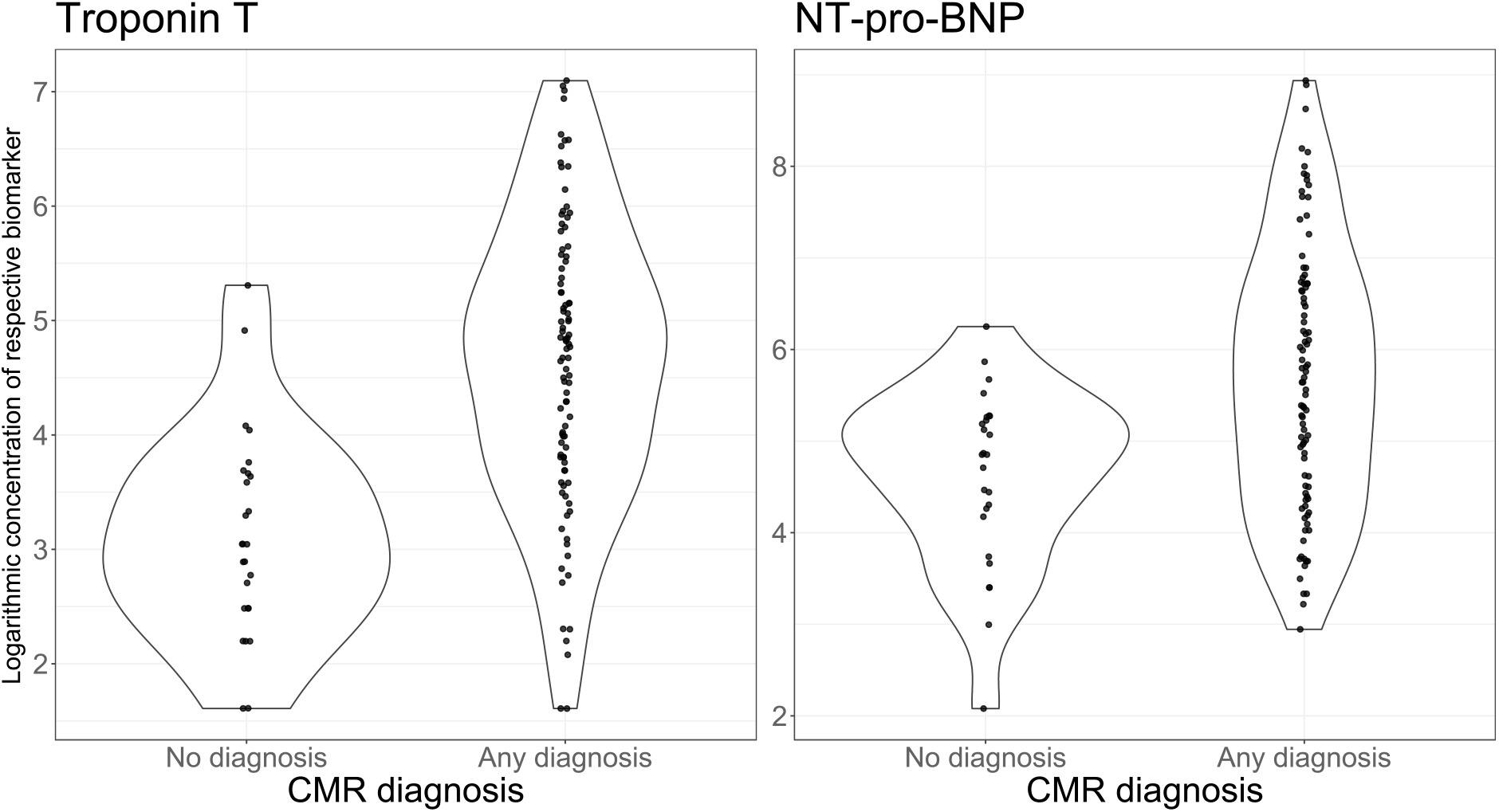
Distributions of troponin T and NT-pro-BNP. Violin plots of the distributions of hs-TnT and NT-pro-BNP levels in patients with normal CMR findings and any diagnosis, respectively. Dots indicate individual data.

**Figure 3.**
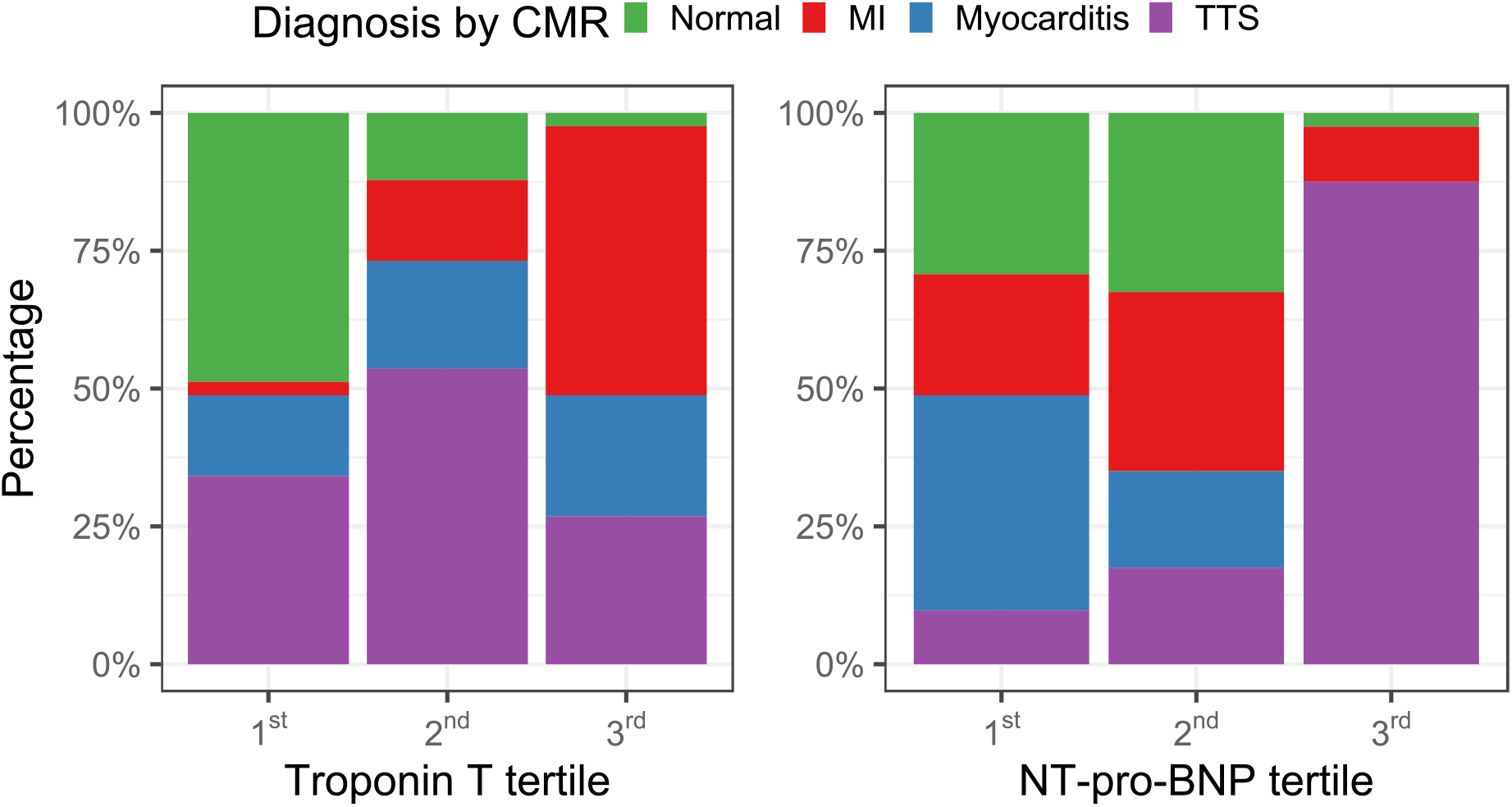
Distribution of CMR diagnosis per tertile of hs-TnT and NT-pro-BNP. Stacked bar chart with bar height indicating prevalence of respective CMR diagnosis per tertile of hs-TnT and NT-pro-BNP. MI – myocardial infarction. TTS – takotsubo syndrome.

**Figure 4.**
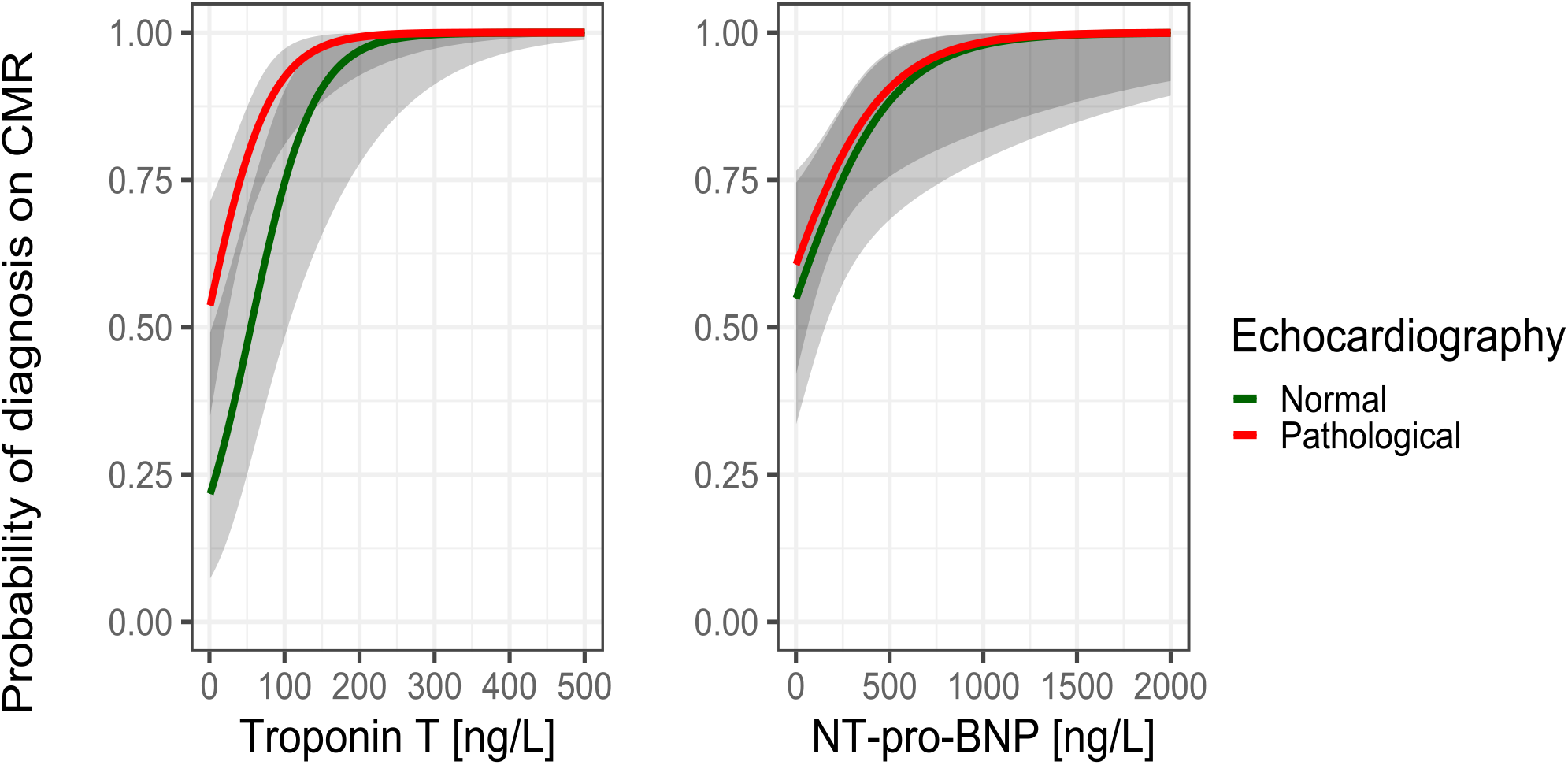
Predicted probabilities of establishing a diagnosis on CMR. Concentrations of hs-TnT and NT-pro-BNP on the x-axes, probabilities of a CMR diagnosis predicted from logistic regression modeling on the y-axes, green lines indicate probabilities for patients with normal echocardiography, red probabilities for patients with pathological echocardiography. Gray shaded areas indicate 95 % confidence intervals of the point estimates.

## Discussion

The main finding of this diagnostic study is that a normal echocardiogram in a patient with MINOCA does not indicate a low probability of obtaining a causal diagnosis with CMR, as indicated by the low specificity and NPV of the criteria used to define a normal echocardiogram. Similarly, even at low levels of hs-TnT and NT-pro-BNP, it is likely that CMR will reveal an underlying diagnosis.

To our knowledge, this is the first study investigating the value of echocardiography in selecting patients for further evaluation with CMR in the setting of MINOCA. The results are in line with the recent findings that the addition of high resolution late gadolinium enhancement to conventional CMR had the strongest impact in patients with normal echocardiography^17^. Also of note, in a post-hoc analysis of the SMINC-2 study, clinical echocardiography identified patients with TTS with a sensitivity and specificity of 57% and 95%, respectively^6^. The results thus illustrate the importance of CMR to image pathological changes of tissue composition such as edema and scarring, even when these changes only lead to minimal or indiscernible alterations in function as determined by echocardiography.

In the current study we found an association between hs-TnT and CMR diagnosis regardless of echocardiography findings. Recently, in a retrospective study of MINOCA patients who had undergone CMR^7^, it was found that the diagnostic yield from CMR was considerably higher in the group of patients with troponin T above the level maximizing the Youden index (sensitivity+specificity-1) for diagnosis. In a larger prospective study of MINOCA patients, it was not possible to identify a troponin level below which < 15% received a diagnosis by CMR^9^. In contrast, in another prospective study, focused on the concordance between clinical and CMR assessment in MINOCA, there were no significant CMR findings in the group of patients with troponin T < 100 ng/L and a normal ECG^18^. The findings of previous studies are in line with the findings from the present prospective study. However, the high yield of CMR even at low levels of hs-TnT, and NT-pro-BNP should be emphasized.

Regarding the patients with normal findings on CMR, it is noteworthy that as all patients included in the present study presented as acute coronary syndromes and all had significantly elevated levels of hs-TnT at inclusion, hence the absence of a causal diagnosis on CMR does not indicate the absence of disease per se. It is likely that the group contains patients with e.g., heart failure with preserved EF and paroxysmal arrythmias, very small Mis and myocarditis cases, as well as TTS with rapid normalization. This is likely reflected in the presence of abnormal echocardiographic findings in this group in the absence of a CMR-based diagnosis. CMR in the present study was performed at a median of 3 days after hospital admission, which increased the diagnostic yield when retrospectively compared to CMR performed later^6^. If an even earlier CMR examination was to be performed, it is not unlikely that even more patients would receive a diagnosis.

It should also be noted that, even if echocardiography cannot readily identify which patients should be further investigated with CMR, the value of echocardiography in assessing biventricular function, valvular heart disease, pulmonary hypertension and signs of pericardial disease in a population with suspected acute coronary syndrome, remains.

### Study limitations

First, the echocardiograms, as well as the blood sampling, were performed on the same day as the CMR examination, 2–4 days after admission. Even though all patients had elevated hs-TnT as a part of their clinical presentation, some had normalized at the time of blood sampling for this study. It is possible that both blood tests and imaging performed at an earlier timepoint would have had a higher accuracy. This is particularly plausible in the case of echocardiography in suspected TTS, where rapid normalization of RWMA often is seen, suggesting that imaging should be performed as early as possible. The optimal time-point for measuring troponin and NT-pro-BNP is not known, and the effects of earlier sampling is unknown in this patient population. Second, the reading of the echocardiographic examinations was done blinded to CMR diagnosis and biomarker levels, but not to the fact that all patients had MINOCA. This might have resulted in false positive assessments of RWMA, which are more easily influenced by subjectivity compared to other parameters. Third, the predicted probabilities for hs-TnT and NT-pro-BNP for identifying patients receiving a diagnosis came from unvalidated models, and should be seen as indicators of trends rather than exact predictions. Finally, the diagnosis for all patients was based on results of the CMR examination, and OCT was not performed. OCT could possibly have identified cases with occult plaque rupture/erosion or spontaneous coronary artery dissection, suggesting a minimal MI, below the ability of detection by CMR, as the underlying cause.

## Conclusion

In patients with MINOCA, a majority will receive a diagnosis by CMR, even in the presence of a normal echocardiogram, or low levels of hs-TnT and NT-pro-BNP. Thus, CMR should be recommended regardless of echocardiographic findings and biomarker levels in the work-up of all patients with MINOCA.

## Data Availability

All data produced in the present study are available upon reasonable request to the authors.

## Abbreviations

CMR: cardiac magnetic resonance imaging
GLS: global longitudinal strain
hs-TnT: high sensitive troponin T
MI: myocardial infarction
MINOCA: myocardial infarction with non-obstructed coronary arteries
NPV: negative predictive value
OCT: optimal coherence tomography
PPV: positive predictive value
RWMA: regional wall motion abnormalities
TTS: takotsubo syndrome

## Acknowledgements

We acknowledge the contributions of the biomedical scientists Margareta Ring PhD, and Maja Kopelev, for performing the echocardiographic investigations.

