## Supplementary material for "Cardiovascular magnetic resonance imaging after normal echocardiography in myocardial infarction with nonobstructed coronary arteries" (from another version of this preprint: version 1, 2022-10-26)

Supplemental material

Logistic regression models for obtaining a CMR diagnosis

Model 1

Specification: CMR diagnosis ~ NT-proBNP + Pathological echo (0/1)

Number of observations: 121

Model likelihood ratio test χ² 18.48, p<0.0001

|  | Coefficient | Standard error | Wald Z | p |
| --- | --- | --- | --- | --- |
| Intercept | 0.1926 | 0.4488 | 0.43 | 0.668 |
| NT-pro-BNP | 0.0037 | 0.0015 | 2.48 | 0.013 |
| Pathological echo | 0.2362 | 0.4888 | 0.48 | 0.629 |

Model 2

Specification: CMR diagnosis ~ Troponin T + Pathological echo (0/1)

Number of observations: 123

Model likelihood ratio test χ² 35.48, p<0.0001

|  | Coefficient | Standard error | Wald Z | p |
| --- | --- | --- | --- | --- |
| Intercept | -1.3058 | 0.6396 | -2.04 | 0.041 |
| Troponin T | 0.0238 | 0.0070 | 3.38 | 0.001 |
| Pathological echo | 1.4312 | 0.5995 | 2.39 | 0.017 |
